# Evaluation of a SARS-CoV-2 rapid antigen test: potential to help reduce community spread?

**DOI:** 10.1101/2020.12.04.20240283

**Authors:** Tuna Toptan, Lisa Eckermann, Annika E. Pfeiffer, Sebastian Hoehl, Sandra Ciesek, Christian Drosten, Victor M. Corman

## Abstract

Severe acute respiratory syndrome coronavirus 2 (SARS-CoV-2) can spread from symptomatic patients with COVID-19, but also from asymptomatic individuals. Therefore, robust surveillance and timely interventions are essential for the control of virus spread within the community. In this regard the frequency of testing and speed of reporting, but not the test sensitivity alone, play a crucial role. In order to reduce the costs and meet the expanding demands in real-time RT-PCR (rRT-PCR) testing for SARS-CoV-2, complementary assays, such as rapid antigen tests, have been developed. Rigorous analysis under varying conditions is required to assess the clinical performance of these tests and to ensure reproducible results. We evaluated the sensitivity and specificity of a recently licensed rapid antigen test using 137 clinical samples in two institutions. Test sensitivity was between 88.2-89.6% when applied to samples with viral loads typically seen in infectious patients. Of 32 rRT-PCR positive samples, 19 demonstrated infectivity in cell culture, and 84% of these samples were reactive with the antigen test. Seven full-genome sequenced SARS-CoV-2 isolates and SARS-CoV-1 were detected with this antigen test, with no cross-reactivity against other common respiratory viruses. Numerous antigen tests are available for SARS-CoV-2 testing and their performance to detect infectious individuals may vary. Head-to-head comparison along with cell culture testing for infectivity may prove useful to identify better performing antigen tests. The antigen test analyzed in this study is easy-to-use, inexpensive, and scalable. It can be helpful in monitoring infection trends and thus has potential to reduce transmission.

## 1. INTRODUCTION

Since the beginning of COVID-19 outbreak in December 2020, the global demand for the Severe acute respiratory syndrome coronavirus 2 (SARS-CoV-2) testing has been steadily increasing. Already back in March 2020, hospitals and laboratories around the world announced their concerns about reagent, consumable material shortages, and limited personal protective equipment. Yet, timely detection and isolation of SARS-CoV-2 infected cases and identification of their contacts are pivotal to slowing down the pandemic.

The main public health strategy during a pandemic relies on robust and easy to perform diagnostic tools that can be used to test large number of samples in a short time. To date the gold standard diagnostic method for SARS-CoV-2 detection [1] is based on real time reverse transcription-PCR (rRT-PCR) technology which has been promptly implemented by the World Health Organization (WHO) [2], Center for Disease Control and Prevention (CDC) [3] protocols, and a number of commercial assays [4]. The SARS-CoV-2 rRT-PCR has high specificity and sensitivity [5, 6]. However, the type and quality of the patient specimen [7, 8], stage of the disease, and the degree of viral replication and/or clearance have an impact on the test outcome [9]. These factors are critical not only for PCR-based but also for other diagnostic test systems aiming to detect the presence of the virus. Hence interpreting a test result for SARS-CoV-2 depends on the accuracy of the test, but the prevalence and the estimated risk of disease before testing should also be taken into consideration.

In many countries SARS-CoV-2 testing is extended to asymptomatic population, e.g. in schools, airports, nursing-homes, and workplaces. This leads to a growing gap between the large number of demand and the laboratory capacities to preform rRT-PCR tests, especially in developing countries. Despite high specificity and sensitivity, rRT-PCR has a disadvantage in point of care testing, because it usually requires professional expertise, expensive reagents and specialized equipment. Therefore, alternative assays, such as rapid antigen detection tests, which can also detect the presence of the virus directly in respiratory samples, have been developed [4] and tested by different groups [10-14]. However, it is vital to determine the sensitivity, specificity of such tests relative to standard rRT-PCR in order to identify the ideal circumstances that their application would be beneficial.

This study was performed to evaluate a novel antigen test produced by R-Biopharm for the detection of SARS-CoV-2 in different specimens and to identify its limitations and potential usage. Different types of materials and verification analysis were used by two institutions independently to assure the reproducibility of the testing and to analyze the potential caveats.

## 2. MATERIALS AND METHODS

### 2.1 Specimen collection

At the Institute of Virology, Charité Berlin stored specimens taken after routine diagnostic were used with no extra procedures required for the study. Cell culture supernantants of respiratory viruses other than SARS-CoV-2 were available at the institute of virology, Charite through a EVD-LabNet EQA (https://www.evd-labnet.eu/; Fischer/Mögling, unpublished data).

At the Institute of Virology, Frankfurt, the clinical samples were collected from subjects as part of registered protocols. Combined oropharyngeal/nasal swabs were collected, stored in 2 ml PBS at 4°C and processed for further analysis within 24 hours.

### 2.2 Cell culture and virus stocks

Caco-2 (human colon carcinoma) were cultured in Minimum Essential Medium (MEM) supplemented with 10% fetal calf serum (FCS). 100 IU/mL of penicillin and 100 g/mL of streptomycin. All culture reagents were purchased from Sigma (St. Louis. MO. USA). The Caco-2 cells were originally obtained from DSMZ (Braunschweig, Germany, no.: ACC 169) differentiated by serial passaging and selected for high permissiveness to virus infection. Caco-2 cells were infected with different viral isolates (FFM1-FFM7) [15] at an MOI 0.1. Cell culture supernatant was harvested 48 h after infection, precleared at 2000 x g for 10 min at room temperature. Aliquots of virus particle containing supernatant were kept at −80°C.

### 2.3 Detection of infectious virus in cell culture

Of the swab-dilution, 500 µL were mixed with 1.5 ml of MEM containing 1% FCS (Sigma-Aldrich; St. Louis, Missouri, USA), 7.5 µg/ml Amphotericin B, and 0.1 mg/ml Primocin, (InvivoGen; San Diego, California, USA). Swab-inoculums were transferred to Caco-2 cells seeded in 5.5 cm2 culture tubes. Cytopathogenic effect (CPE) was assessed daily for up to seven days or until cell lysis occurred.

### 2.4 Rapid Antigen Test

Rapid antigen test was provided by R-Biopharm. Test was performed according to the manufacturer’s recommendations and evaluated visually by four or six-eye principle. Briefly, samples were vortexed for 20 sec. 50 µl from Solution A (blue) and B (yellow) were dispensed in clean 1.5 ml reaction tubes which leads to green coloring. Immediately 50 µl of the test samples were added to the reaction mixture. Samples were then mixed briefly and incubated for 10 min at room temperature. Test strips were placed in to mixture vertically to allow absorption. Test results were evaluated after 10 min. Intensities of the test bands were compared to control band categorized as follows: +++ (test band intensity stronger than the control), ++ (test and control bans intensity are similar), + (test band intensity is weaker than the control). Antigen testing for viable SARS-CoV-2 and SARS-CoV-1 cell culture supernatants was performed in a BSL-3 laboratory.

### 2.5 RNA extraction and rRT-PCR analysis

At the Institute of Virology, Charité Berlin, stored samples (swab resuspended in 1.5 mL of phosphate-buffered saline) were anonymized before testing. After thawing at RT all samples were analyzed by antigen test and rRT-PCR in parallel. RNA extraction for rRT-PCR was done by using the MagNA Pure 96 system, using 100 μl of sample, eluted in 100 μl. rRT-PCR was done as published previously [1].

At the Institute of Virology in Frankfurt the SARS-CoV-2 test (Cobas, Roche, Basel, Switzerland) was performed on the rRT-PCR automated Cobas 6800 system. Of the swab-dilution, 1000 µl aliquots were mixed with lysis buffer (1:1 ratio) and 500 μL aliquots were transferred to barcoded secondary tubes, loaded on the Cobas 6800 system, and tested with Cobas SARS-CoV-2 master mix containing an internal RNA control and primer-probe sets towards ORF1 and E-gene according to the manufacturer’s instructions.

Within seven days of virus inoculation using clinical sample material, culture supernatant was collected to perform rRT-PCR in order to confirm productive virus replication. RNA was isolated from 100 µL cell culture supernatant using the QIAcube HT instrument and QIAamp 96 Virus QIAcube HT Kit (Qiagen; Hilden, Germany) according to the manufacturer’s instructions. SARS-CoV-2 RNA was analyzed by rRT-PCR using the Luna Universal One-Step RT-qPCR Kit (New England Biolabs; Ipswich, Massachusetts, USA) and primers targeting RNA-dependent RNA polymerase (RdRp) [15]. RdRP_SARSr-F2 (GTGARATGGTCATGTGTGGCGG)RdRP_SARSr-R1(CARATGTTAAASACACTATTAGCATA).

### 2.6 Statistical Analysis

The number of positive samples were compared two by two contingency table. The agreement between the antigen test and rRT-PCR techniques was evaluated using the Cohen’s weighted kappa index (K value) [16]. K value interpretations were categorized as follows: <0.20 is poor, 0.21-0.40 is fair, 0.41-0.60 is moderate agreement, 0.61-0.80 is substantial agreement and 0.81-1.00 is almost perfect agreement [17].

### 2.7 Ethical Statement

The use of stored clinical samples for validation of diagnostic methods without person related data is covered by section 25 of the Berlin hospital law and does not require ethical or legal clearance. The use of anonymized clinical samples for validation of diagnostic methods does not require ethical clearance by the Goethe University, Frankfurt.

## 3. RESULTS

Rapid antigen test sensitivity and specificity were evaluated by two independent institutions using various number of clinical samples. rRT-PCR was used as a reference test system. We deemed individuals to be uninfected with SARS-CoV-2 when a negative result was obtained by rRT-PCR.

In the Institute of Virology, Charité, Berlin, a total of 67 stored patient samples were available for the study. Of these, 58 were rRT-PCR positive with cycle threshold (cT) range between 18.77-40 corresponding to 2.5×10^9^ −1380 RNA genome copies/ml (**Table S1**), representing 86.6% (58/67) of the clinical samples analyzed (**Figure 1A**). When the rRT-PCR results were used as a reference, the antigen test diagnosed SARS-CoV-2 infection status with a sensitivity of 77.6% (45/58) and a specificity of 100% (9/9) (**Table 1**). After re-evaluating the data based on the acceptable analytic sensitivity and limit of detection suggested by WHO [18], we identified 48 samples with ≥10^6^ RNA genome copies/ml. Rapid antigen test performed with 89.6% sensitivity for this sample set (**Table S1**). Of these, 40 samples had ∼2.23×10^6^ or more RNA genome copies/ml and reacted positive with the antigen test (**Table 1**). In contrast samples with less than 7.63×10^5^ RNA copies/ml were negative (**Figure 1A, Table S1**). Cohen’s weighted kappa value of 0.482 indicated moderate agreement between the rRT-PCR and the rapid antigen test (**Table 2**). The overall concordance between the rRT-PCR and the antigen test was 80.6% (54/67).

**Table 1.**
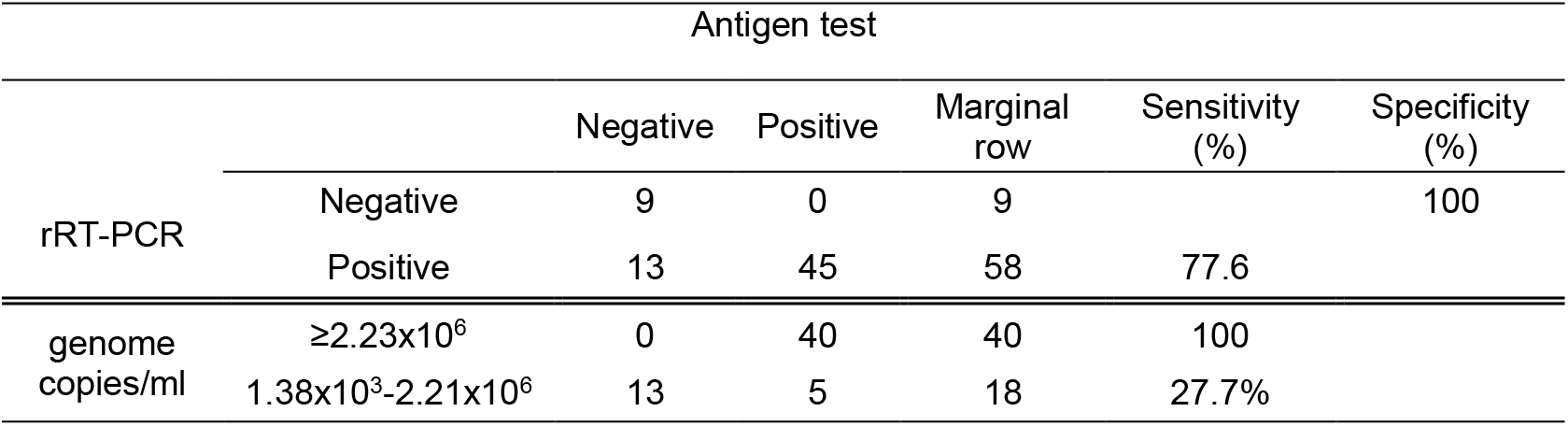
Sensitivity and specificity of the antigen detection test in comparison to rRT-PCR.

**Table 2.**
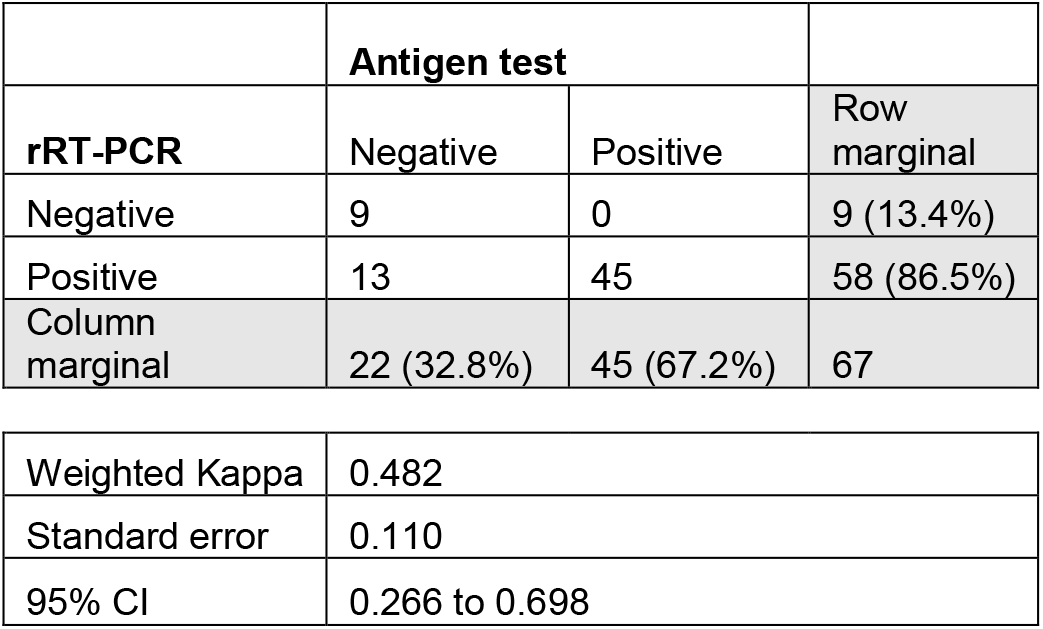
Cohen’s weighted kappa coefficient between rapid antigen test and rRT-PCR.

|  | Antigen test |  |  |
| --- | --- | --- | --- |
| rRT-PCR | Negative | Positive | Row marginal |
| Negative | 9 | 0 | 9 (13.4%) |
| Positive | 13 | 45 | 58 (86.5%) |
| Column marginal | 22 (32.8%) | 45 (67.2%) | 67 |

|  |  |
| --- | --- |
| Weighted Kappa | 0.482 |
| Standard error | 0.110 |
| 95% CI | 0.266 to 0.698 |

**Figure 1.**
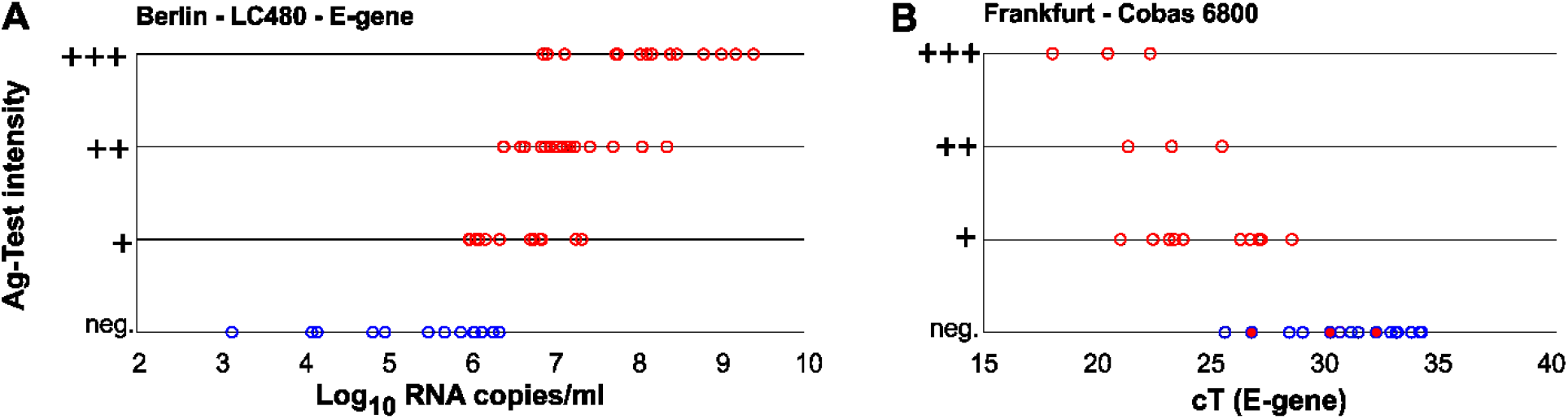
Antigen test analysis performed in Berlin (A) and Frankfurt (B). **A**. Log_10_ RNA copies/ml and corresponding antigen (Ag) detection test results (red circles positive n: 45, blue circles negative n: 13) intensity for each rRT-PCR positive sample (n: 58). **B**. Cycle threshold (cT) value and corresponding antigen (Ag) detection test results (red circles positive n: 16, blue circles negative n: 16) intensity for each rRT-PCR positive sample (n: 32). 32 rRT-PCR positive samples were tested in cell culture for infectivity. All Ag-test positive (n:16, red circles) and three Ag-test negative (red-filled blue circles) samples displayed CPEs after inoculating in Caco-2 cells (**Table S2**). Intensities of the test bands were compared to control band and designated as follows: +++ (test band intensity stronger than the control), ++ (test and control bans intensity are similar), + (test band intensity is weaker than the control).

Certain rapid tests may be used at the point-of-care and thus offer benefits for the detection and management of infectious diseases. In order to assess the potential of the rapid antigen test in this context, 70 nasopharyngeal samples freshly collected from individuals living in a shared housing were analyzed head to head by rRT-PCR using Cobas 6800 system, rapid antigen test, and cell culture using Caco-2 cells to determine the infectivity (Institute of Medical Virology, Goethe University, Frankfurt). 45.7% (32/70) of the clinical samples were diagnosed positive for SARS-CoV-2 by rRT-PCR with cT values ranging between 18.01-35.98 (**Figure 1B, Table S2**). The antigen test diagnosed the infection status with a sensitivity of 50% (16/32) and a specificity of 100% (**Table 3**). Re-evaluating the data based on the limit of detection, sensitivity was determined to be 88.2% for samples with cT values <28, and it was reduced in the group of samples with cT values ≥ 28 (6.7%) (**Table 3**). Cohen’s weighted kappa value of 0.521 indicated moderate agreement between rRT-PCR and the rapid antigen test (**Table 4**). The overall concordance between the rRT-PCR and the antigen test was 77.1% (54/70) (**Table 4**).

**Table 3.**
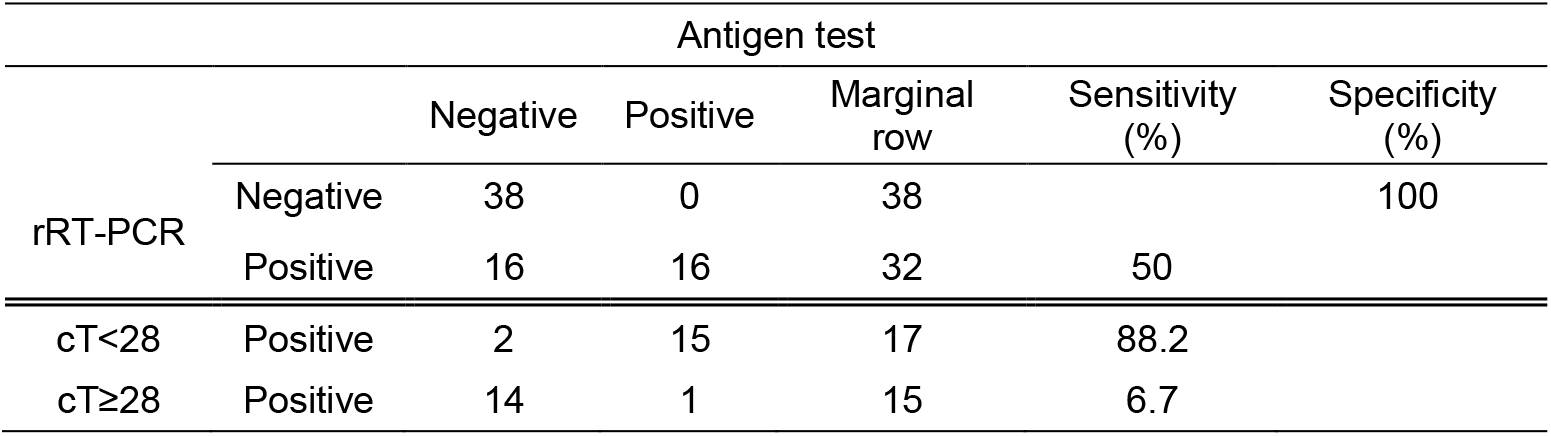
Comparison of the clinical diagnostic performance of rapid antigen test with rRT-PCR.

**Table 4.**
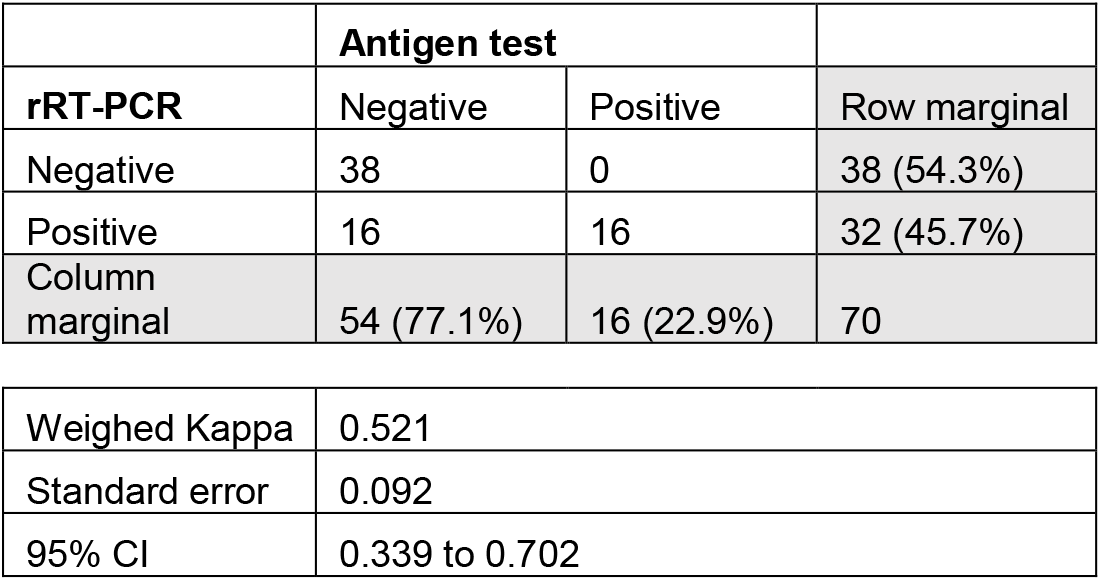
Cohen’s weighted kappa coefficient between rapid antigen test and rRT-PCR.

|  | Antigen test |  |  |
| --- | --- | --- | --- |
| rRT-PCR | Negative | Positive | Row marginal |
| Negative | 38 | 0 | 38 (54.3%) |
| Positive | 16 | 16 | 32 (45.7%) |
| Column marginal | 54 (77.1%) | 16 (22.9%) | 70 |

|  |  |
| --- | --- |
| Weighed Kappa | 0.521 |
| Standard error | 0.092 |
| 95% CI | 0.339 to 0.702 |

rRT-PCR is a highly sensitive method to detect viral RNA molecules from clinical samples. However, viral RNA can persist in different body parts and can be detected in specimens for much longer than the presence of viable virus [19]. Thus demonstration of infectivity on permissive cell lines *in vitro* is a more reliable surrogate for infectivity and virus transmission. Therefore, we attempted virus isolation by inoculating rRT-PCR positive samples in Caco-2 cells. Cytopathic changes were monitored daily by microscopy for a week and subsequently aliquots of culture supernatant were tested to verify viral RNA copies (**Table S2**). For samples that are positive for both antigen test and rRT-PCR (16/32, cT 18.01-28.45), we observed cytopathic effects (CPE) in cell culture 1-3 days after inoculation (**Figure 1B, Table S2**). Three samples that had a negative result in the antigen test, but were positive by rRT-PCR (cT values 26.69, 30.12, and 32.13) displayed CPE as well. Other 13 antigen-test negative samples with higher cT values (indicating lower viral load) between 28.34-34.12 were not infectious in cell culture. Interestingly, one sample with a relatively low cT value 25.53, did not show any CPE in cell culture and was also negative for the antigen test (**Table S2**).

In order to investigate potential cross reactivity among common coronaviruses and other respiratory viruses, infectious and heat inactivated (4 h at 60°C) cell culture supernatants were tested (**Table 5**). SARS-CoV-1 and SARS-CoV-2 tested positive with the antigen test, as expected. The antigen test did not display any cross-reactivity with the other respiratory and endemic corona viruses listed in **Table 5**.

**Table 5.**
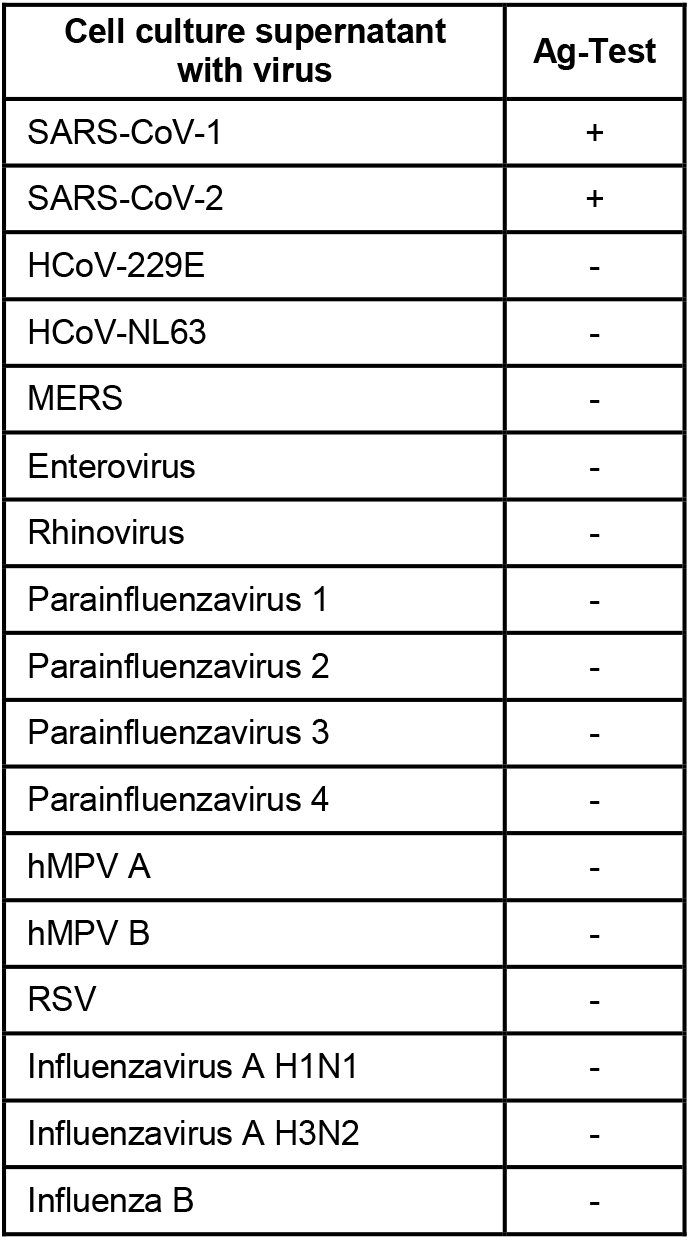
Rapid Antigen Test results using different respiratory virus cell culture supernatant stocks.

We further evaluated the detection sensitivity among different SARS-CoV-2 isolates. Here we used cell culture supernatant collected from Caco-2 cells infected with seven different isolates [15] and SARS-CoV-1 (**Figure 3**). The virus stocks were thawed at room temperature and a total of six 10-fold dilutions were prepared in PBS. The antigen test was performed and evaluated immediately (**Figure 3A**). In parallel, aliquots of the dilutions were mixed with lysis buffer used for RNA extraction to inactivate the virus. rRT-PCR was performed for two different gene targets ORF1 and E-gene that resulted in similar cT values (**Figure 3B, Table S3**). 10-fold serial dilutions led to ∼3 cT difference in rRT-PCR for each set as anticipated. According to our results the limit of detection was between 100-560 RNA copies/ml which is in line with the manufacturer’s findings. We previously identified RG203KR mutations in FFM3, FFM4 and FFM6 and S→L mutation in FFM1 within the nucleocapsid protein coding region [15]. According to GISAID classification the GR clade, carrying the combination of Spike D614G and nucleocapsid RG203KR mutations, is currently the most common representative of the SARS-CoV-2 population worldwide [20]. Our results suggest that the presence of the RG203KR mutation did not interfere with the antigen test performance.

**Figure 3.**
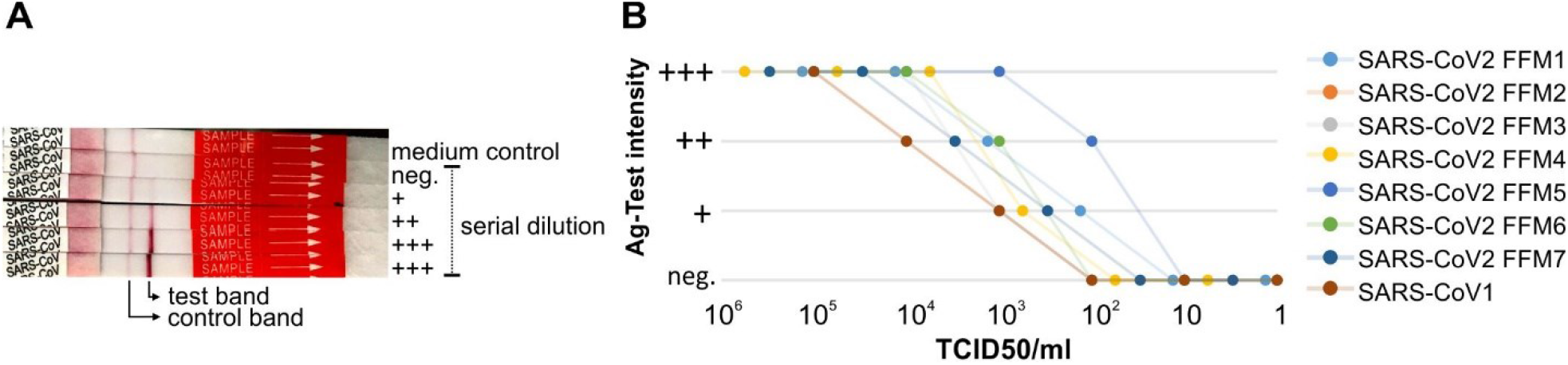
Rapid Antigen Test Results for SARS-CoV-1 and SARS-CoV-2 isolates. **A**. Representative lateral flow assay using serially diluted virus stock. Intensities of the test bands were compared to control band and designated as follows: +++ (test band intensity stronger than the control), ++ (test and control bans intensity are similar), + (test band intensity is weaker than the control). **B**. TCID50/ml values and corresponding antigen (Ag) detection test intensity for serially diluted SARS-CoV-2 isolates FFM1-7 and SARS-CoV-1 are shown. Representative result of two experiments.

## 4. DISCUSSION

In this study we validated the assay performance of a recently approved rapid antigen test in two independent institutions using a total of 137 clinical samples. Although the test specificity was 100% for this particular sample set, overall sensitivity was low (50-77.6%), yet re-analyzing samples with higher viral loads showed good correlation (88.2-89.6%). Previous studies reported that lower cT values are associated with higher viral culture positivity [21, 22]. There is currently no direct evidence whether cell culture positivity or higher viral load correlates with contagiousness of an individual, however, it is commonly recognized as the surrogate of infectivity [23]. Since an important aspect of using point-of-care testing is to able to identify infected individuals who are infectious and can potentially transmit the virus, we performed correlation analysis within a group of clinical samples tested. 19 out of 32 SARS-CoV-2 infected individuals were positive in cell culture. The antigen test detected 16 out of 19 these (84%). In contrast 43.7% (14/32) of the samples were not infectious in cell culture, yet positive by rRT-PCR, probably due to persisting genomic and subgenomic viral RNA within the collected sample. We detected an excess amount of viral RNA in cell culture supernatants due to high replication capacity of the virus in permissive cells, despite a negative antigen test result. This might explain the cT discrepancy between the cell culture supernatant and clinical samples. Limited clinical sample size is the major limitation of this study. Future efforts should aim to monitor frequent sampling of larger groups and to compare different rapid antigen tests, different sampling sites along with infectivity correlation in cell culture.

Our results suggest that the rapid antigen test can detect SARS-CoV-2 infected individuals with high viral loads and has potential in determining highly contagious individuals. Despite low analytic sensitivity, rapid antigen tests are inexpensive and therefore can be used frequently for detecting infected individuals who are asymptomatic, pre-symptomatic and without known or suspected exposure to SARS-CoV-2 [24]. They can be beneficial in congregate settings, such as a long-term care facility or a correctional facility, workplace, or a school testing its students, faculty, and staff. Rapid antigen tests probably perform best during the early stages of SARS-CoV-2 infection when the viral load is higher.

## Data Availability

No external datasets or supplementary material online at other repositories.

## ACKNOWLEDGMENTS

Parts of this work was funded by the German Ministry of Health (Konsiliarlabor für Coronaviren) to C.D. and V.M.C. and by the German Ministry of Research through projects VARIPath (01KI2021) to V.M.C.

This project was funded in part by the German Federal Ministry of Education and Research (Bundesministerium für Bildung und Forschung, BMBF) (NaFoUniMedCovid19 – B-FAST, EVIPAN, FKZ: 01KX2021).

We thank Lara Jeworowski and Tobias Bleicker for their assistance, Felix Drexler for providing heat inactivated respiratory viruses for specificity testing, Dr. Holger Rabenau for organizational support, Marhild Kortenbusch and Regine Jeck for technical assistance.

## SUPPLEMENTARY TABLES

**Table S1.** Summary of rRT-PCR and antigen testing using clinical samples (Charite, Berlin). neg: negative antigen test. N/D: not detectable.

| Sample ID | rRT-PCR |  | Antigen test |
| --- | --- | --- | --- |
|  | E-gene | Log10 RNA copies/mL | Intensity |
| #65 | 18,77 | 9,40 | +++ |
| #19 | 19,48 | 9,18 | +++ |
| #60 | 20,02 | 9,01 | +++ |
| #56 | 20,72 | 8,80 | +++ |
| #22 | 21,78 | 8,47 | +++ |
| #66 | 22,01 | 8,40 | +++ |
| #25 | 22,02 | 8,40 | +++ |
| #67 | 22,16 | 8,35 | ++ |
| #21 | 22,74 | 8,17 | +++ |
| #20 | 22,94 | 8,11 | +++ |
| #57 | 23,11 | 8,06 | ++ |
| #15 | 23,19 | 8,04 | +++ |
| #53 | 24,07 | 7,76 | +++ |
| #26 | 24,15 | 7,74 | +++ |
| #8 | 24,24 | 7,71 | ++ |
| #12 | 25,15 | 7,43 | ++ |
| #27 | 25,47 | 7,33 | + |
| #58 | 25,7 | 7,26 | + |
| #14 | 25,74 | 7,25 | ++ |
| #1 | 25,92 | 7,19 | ++ |
| #18 | 26,04 | 7,16 | ++ |
| #64 | 26,12 | 7,13 | +++ |
| #4 | 26,14 | 7,13 | +++ |
| #10 | 26,22 | 7,10 | ++ |
| #11 | 26,44 | 7,03 | ++ |
| #23 | 26,71 | 6,95 | ++ |
| #3 | 26,81 | 6,92 | +++ |
| #28 | 26,89 | 6,90 | ++ |
| #29 | 26,89 | 6,90 | ++ |
| #61 | 26,98 | 6,87 | +++ |
| #5 | 27,03 | 6,85 | + |
| #13 | 27,05 | 6,85 | ++ |
| #59 | 27,09 | 6,83 | + |
| #55 | 27,32 | 6,76 | + |
| #49 | 27,47 | 6,72 | + |
| #2 | 27,69 | 6,65 | ++ |
| #17 | 27,85 | 6,60 | ++ |
| #6 | 28,49 | 6,40 | ++ |
| #7 | 28,52 | 6,39 | ++ |
| #9 | 28,67 | 6,35 | + |
| #46 | 28,68 | 6,34 | neg |
| #50 | 28,92 | 6,27 | neg |
| #51 | 29,21 | 6,18 | + |
| #47 | 29,36 | 6,14 | neg |
| #24 | 29,46 | 6,10 | + |
| #32 | 29,59 | 6,06 | + |
| #33 | 29,64 | 6,05 | neg |
| #31 | 29,68 | 6,04 | neg |
| #16 | 29,83 | 5,99 | + |
| #54 | 29,87 | 5,98 | + |
| #52 | 30,18 | 5,88 | neg |
| #48 | 30,81 | 5,69 | neg |
| #30 | 31,45 | 5,49 | neg |
| #39 | 33,13 | 4,97 | neg |
| #43 | 33,61 | 4,83 | neg |
| #62 | 35,76 | 4,16 | neg |
| #63 | 35,98 | 4,10 | neg |
| #41 | 40 | 3,14 | neg |
| #34 | N/D | N/D | neg |
| #35 | N/D | N/D | neg |
| #36 | N/D | N/D | neg |
| #37 | N/D | N/D | neg |
| #38 | N/D | N/D | neg |
| #40 | N/D | N/D | neg |
| #42 | N/D | N/D | neg |
| #44 | N/D | N/D | neg |
| #45 | N/D | N/D | neg |

**Table S2.**
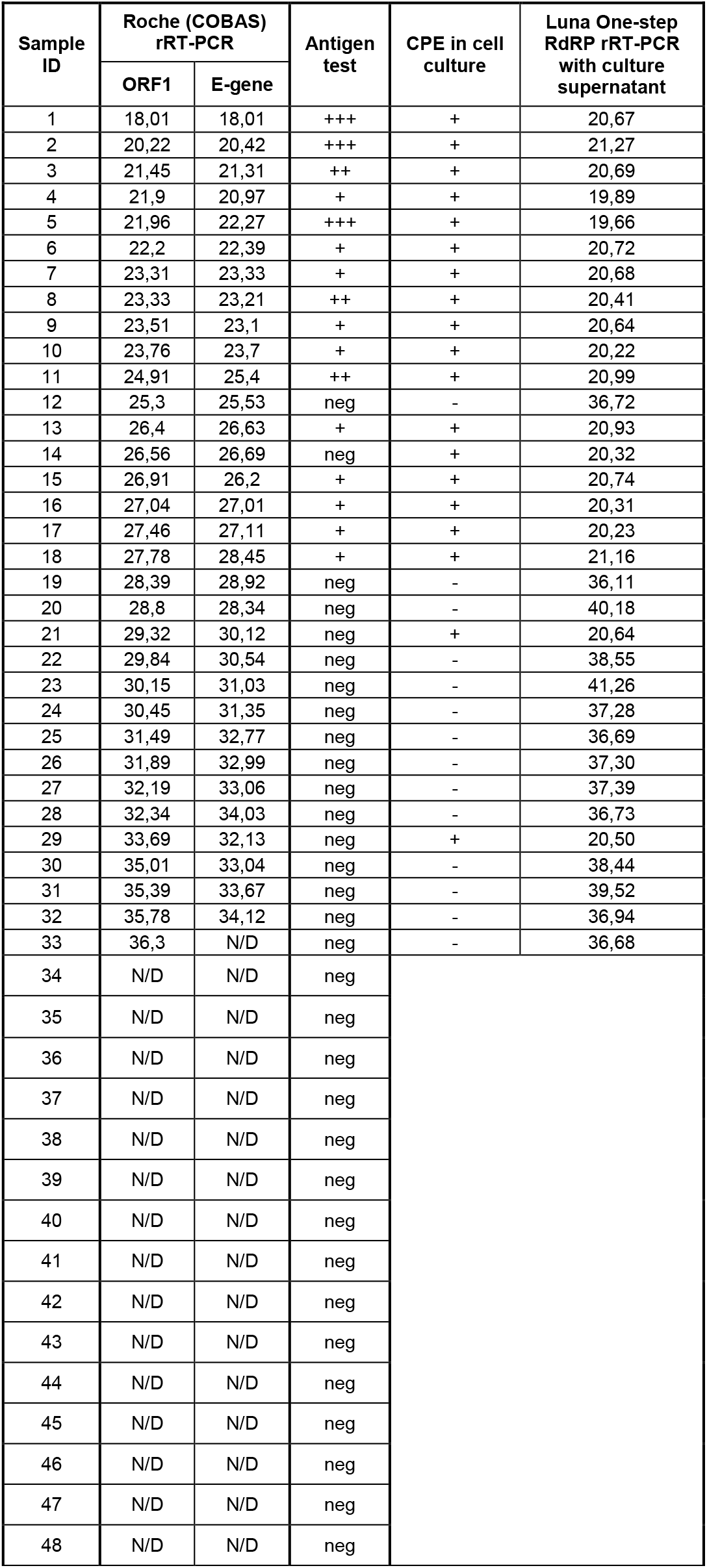
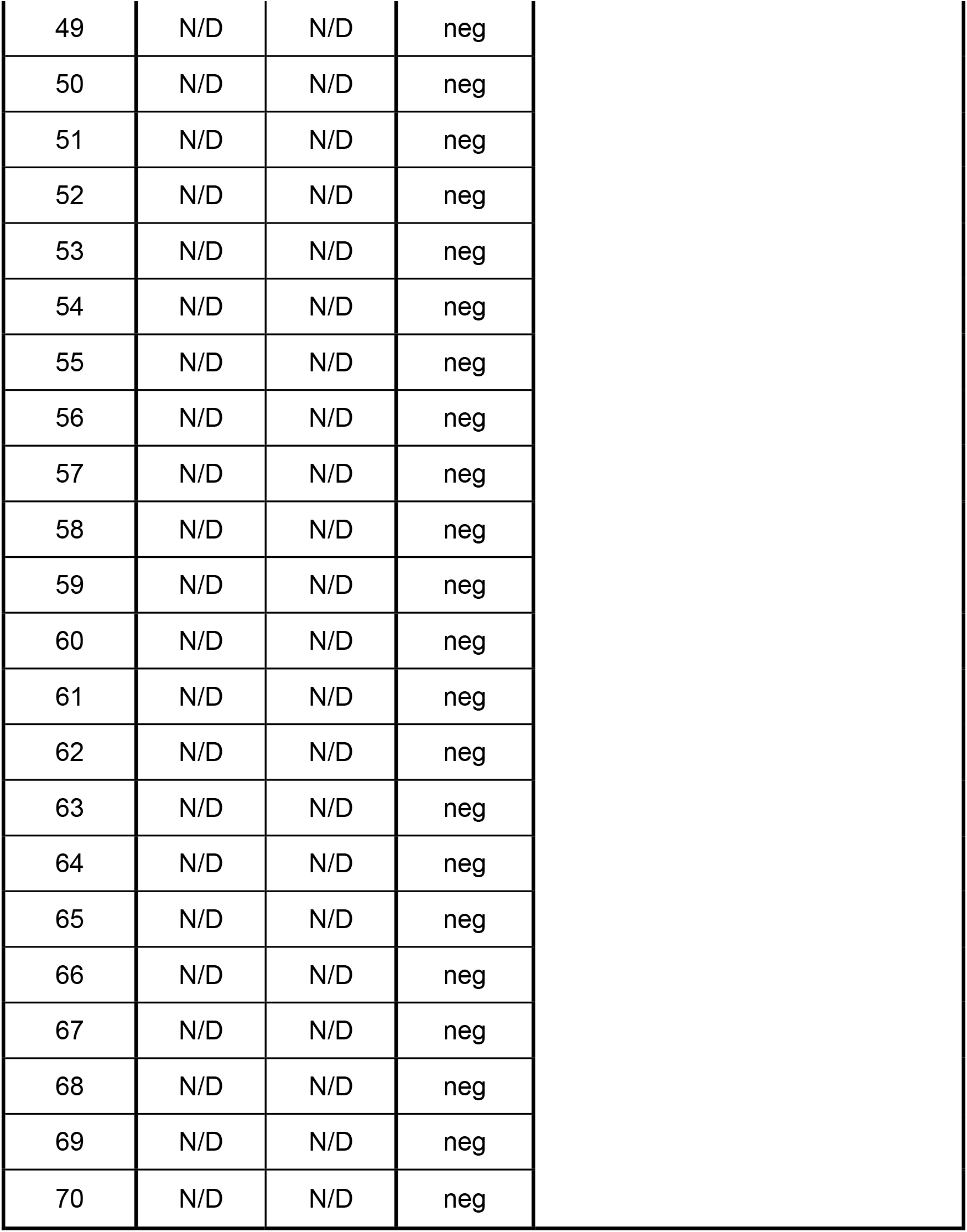
Summary of rRT-PCR (Cobas), antigen testing, virus isolation in cell culture, rRT-PCR with cell culture supernatant of 70 clinical samples (University Hospital Frankfurt). neg: negative antigen test. N/D: not detectable.

| Sample ID | Roche (COBAS) rRT-PCR |  | Antigen test | CPE in cell culture | Luna One-step RdRP rRT-PCR with culture supernatant |
| --- | --- | --- | --- | --- | --- |
|  | ORF1 | E-gene |  |  |  |
| 1 | 18,01 | 18,01 | +++ | + | 20,67 |
| 2 | 20,22 | 20,42 | +++ | + | 21,27 |
| 3 | 21,45 | 21,31 | ++ | + | 20,69 |
| 4 | 21,9 | 20,97 | + | + | 19,89 |
| 5 | 21,96 | 22,27 | +++ | + | 19,66 |
| 6 | 22,2 | 22,39 | + | + | 20,72 |
| 7 | 23,31 | 23,33 | + | + | 20,68 |
| 8 | 23,33 | 23,21 | ++ | + | 20,41 |
| 9 | 23,51 | 23,1 | + | + | 20,64 |
| 10 | 23,76 | 23,7 | + | + | 20,22 |
| 11 | 24,91 | 25,4 | ++ | + | 20,99 |
| 12 | 25,3 | 25,53 | neg | - | 36,72 |
| 13 | 26,4 | 26,63 | + | + | 20,93 |
| 14 | 26,56 | 26,69 | neg | + | 20,32 |
| 15 | 26,91 | 26,2 | + | + | 20,74 |
| 16 | 27,04 | 27,01 | + | + | 20,31 |
| 17 | 27,46 | 27,11 | + | + | 20,23 |
| 18 | 27,78 | 28,45 | + | + | 21,16 |
| 19 | 28,39 | 28,92 | neg | - | 36,11 |
| 20 | 28,8 | 28,34 | neg | - | 40,18 |
| 21 | 29,32 | 30,12 | neg | + | 20,64 |
| 22 | 29,84 | 30,54 | neg | - | 38,55 |
| 23 | 30,15 | 31,03 | neg | - | 41,26 |
| 24 | 30,45 | 31,35 | neg | - | 37,28 |
| 25 | 31,49 | 32,77 | neg | - | 36,69 |
| 26 | 31,89 | 32,99 | neg | - | 37,30 |
| 27 | 32,19 | 33,06 | neg | - | 37,39 |
| 28 | 32,34 | 34,03 | neg | - | 36,73 |
| 29 | 33,69 | 32,13 | neg | + | 20,50 |
| 30 | 35,01 | 33,04 | neg | - | 38,44 |
| 31 | 35,39 | 33,67 | neg | - | 39,52 |
| 32 | 35,78 | 34,12 | neg | - | 36,94 |
| 33 | 36,3 | N/D | neg | - | 36,68 |
| 34 | N/D | N/D | neg |  |  |
| 35 | N/D | N/D | neg |  |  |
| 36 | N/D | N/D | neg |  |  |
| 37 | N/D | N/D | neg |  |  |
| 38 | N/D | N/D | neg |  |  |
| 39 | N/D | N/D | neg |  |  |
| 40 | N/D | N/D | neg |  |  |
| 41 | N/D | N/D | neg |  |  |
| 42 | N/D | N/D | neg |  |  |
| 43 | N/D | N/D | neg |  |  |
| 44 | N/D | N/D | neg |  |  |
| 45 | N/D | N/D | neg |  |  |
| 46 | N/D | N/D | neg |  |  |
| 47 | N/D | N/D | neg |  |  |
| 48 | N/D | N/D | neg |  |  |

|  |  |  |  |
| --- | --- | --- | --- |
| 49 | N/D | N/D | neg |
| 50 | N/D | N/D | neg |
| 51 | N/D | N/D | neg |
| 52 | N/D | N/D | neg |
| 53 | N/D | N/D | neg |
| 54 | N/D | N/D | neg |
| 55 | N/D | N/D | neg |
| 56 | N/D | N/D | neg |
| 57 | N/D | N/D | neg |
| 58 | N/D | N/D | neg |
| 59 | N/D | N/D | neg |
| 60 | N/D | N/D | neg |
| 61 | N/D | N/D | neg |
| 62 | N/D | N/D | neg |
| 63 | N/D | N/D | neg |
| 64 | N/D | N/D | neg |
| 65 | N/D | N/D | neg |
| 66 | N/D | N/D | neg |
| 67 | N/D | N/D | neg |
| 68 | N/D | N/D | neg |
| 69 | N/D | N/D | neg |
| 70 | N/D | N/D | neg |

**Table S3.** Cycle threshold (cT) value for ORF1 and E-gene rRT-PCR and rapid antigen test results for serially diluted SARS-CoV-2 isolates and SARS-CoV-1. neg: negative antigen test.

| SAMPLE | TCID<sub>50</sub>/ml | rRT-PCR (Cobas) |  | Ag-Test |
| --- | --- | --- | --- | --- |
|  |  | ORF1 | E-gene | Intensity |
| SARS-CoV-2<br>FFM1 | 1,3E+05 | 13,12 | 13,86 | +++ |
|  | 1,3E+04 | 16,56 | 16,79 | +++ |
|  | 1,3E+03 | 20,22 | 20,52 | ++ |
|  | 1,3E+02 | 23,86 | 24,10 | + |
|  | 1,3E+01 | 27,24 | 27,63 | neg |
|  | 1,3E+00 | 30,50 | 31,02 | neg |
| SARS-CoV-2<br>FFM2 | 1,0E+05 | 20,12 | 20,37 | +++ |
|  | 1,0E+04 | 23,23 | 23,62 | ++ |
|  | 1,0E+03 | 27,07 | 27,29 | + |
|  | 1,0E+02 | 30,25 | 30,53 | neg |
|  | 1,0E+01 | 33,24 | 33,83 | neg |
|  | 1,0E+00 | 34,78 | 36,23 | neg |
| SARS-CoV-2<br>FFM3 | 1,0E+05 | 16,36 | 16,63 | +++ |
|  | 1,0E+04 | 19,78 | 19,95 | +++ |
|  | 1,0E+03 | 23,45 | 23,54 | + |
|  | 1,0E+02 | 26,72 | 27,14 | neg |
|  | 1,0E+01 | 30,15 | 30,68 | neg |
|  | 1,0E+00 | 33,17 | 33,87 | neg |
| SARS-CoV-2<br>FFM4 | 5,6E+05 | 12,7 | 13,4 | +++ |
|  | 5,6E+04 | 16,3 | 16,78 | +++ |
|  | 5,6E+03 | 19,46 | 19,73 | +++ |
|  | 5,6E+02 | 23,3 | 23,5 | + |
|  | 5,6E+01 | 26,61 | 26,93 | neg |
|  | 5,6E+00 | 29,7 | 30 | neg |
| SARS-CoV-2<br>FFM5 | 1,0E+05 | 13,96 | 14,08 | +++ |
|  | 1,0E+04 | 16,66 | 16,81 | +++ |
|  | 1,0E+03 | 20,13 | 20,33 | +++ |
|  | 1,0E+02 | 23,26 | 23,64 | ++ |
|  | 1,0E+01 | 27,01 | 27,08 | + |
|  | 1,0E+00 | 29,74 | 30,11 | neg |
| SARS-CoV-2<br>FFM6 | 1,0E+05 | 16,1 | 16,35 | +++ |
|  | 1,0E+04 | 19,57 | 19,83 | +++ |
|  | 1,0E+03 | 23,43 | 23,47 | ++ |
|  | 1,0E+02 | 26,36 | 26,63 | + |
|  | 1,0E+01 | 30,04 | 30,64 | neg |
|  | 1,0E+00 | 33,36 | 34,05 | neg |
| SARS-CoV-2<br>FFM7 | 3,0E+05 | 14,87 | 15,12 | +++ |
|  | 3,0E+04 | 17,71 | 18,02 | +++ |
|  | 3,0E+03 | 21,95 | 22,4 | ++ |
|  | 3,0E+02 | 25,4 | 26,12 | + |
|  | 3,0E+01 | 28,81 | 29,26 | neg |
|  | 3,0E+00 | 32,11 | 32,72 | neg |
| SARS-CoV-1 | 1,0E+05 | n/d | 17 | +++ |
|  | 1,0E+04 | n/d | 20,2 | ++ |
|  | 1,0E+03 | n/d | 24,54 | + |
|  | 1,0E+02 | n/d | 27,71 | neg |
|  | 1,0E+01 | n/d | 31,1 | neg |
|  | 1,0E+00 | n/d | 34,21 | neg |

